# COVID-19 and Mental Health: Predicted Mental Health Status is Associated with Clinical Symptoms and Pandemic-Related Psychological and Behavioral Responses

**DOI:** 10.1101/2021.10.12.21264902

**Authors:** Joyce Y. Chung, Alison Gibbons, Lauren Atlas, Elizabeth Ballard, Monique Ernst, Shruti Japee, Cristan Farmer, Jacob Shaw, Francisco Pereira

**Affiliations:** National Institute of Mental Health, 10 Center Drive, NIH Building 10, Room 6-5340, Bethesda, MD 20892, USA

**Keywords:** COVID-19, Pandemic, Mental health status, Prediction, Mental health outcomes

## Abstract

**Background:** The COVID-19 pandemic led to dramatic threats to health and social life. Study objectives - develop a prediction model leveraging subsample of known Patient/Controls and evaluate the relationship of predicted mental health status to clinical outcome measures and pandemic-related psychological and behavioral responses during lockdown (spring/summer 2020).

**Methods:** Online cohort study conducted by National Institute of Mental Health Intramural Research Program. Convenience sample of English-speaking adults (enrolled 4/4–5/16/20; n=1,992). Enrollment measures: demographics, clinical history, functional status, psychiatric and family history, alcohol/drug use. Outcome measures (enrollment and q2 weeks/6 months): distress, loneliness, mental health symptoms, and COVID-19 survey. NIMH IRP Patient/Controls survey responses informed assignment of Patient Probability Scores (PPS) for all participants. Regression models analyzed the relationship between PPS and outcome measures.

**Outcomes:** Mean age 46.0 (±14.7), female (82.4%), white (88.9 %). PPS correlated with distress, loneliness, depression, and mental health factors. PPS associated with negative psychological responses to COVID-19. Worry about mental health (OR 1.46) exceeded worry about physical health (OR 1.13). PPS not associated with adherence to social distancing guidelines but was with stress related to social distancing and worries about infection of self/others.

**Interpretation:** Mental health status (PPS) was associated with concurrent clinical ratings and COVID-specific negative responses. A focus on mental health during the pandemic is warranted, especially among those with mental health vulnerabilities. We will include PPS when conducting longitudinal analyses of mental health trajectories and risk and resilience factors that may account for differing clinical outcomes.

**Funding:** NIMH (ZIAMH002922); NCCIH (ZIAAT000030)

## BACKGROUND

The COVID-19 pandemic has led to dramatic changes in public health, social life, and economic stability, all of which can have widespread behavioral health impacts.^1-3^ Estimates during early spring 2020 found a three-fold increase in depressive symptoms (27.8%) compared to the past (8.5%) in a nationally representative population-based cohort.^4^ This finding was mirrored by data collected by the U.S. Census Bureau Household Pulse and National Health Interview Survey in a similar period which found a three-fold rate of screening positive for anxiety disorder, depressive disorder, or both compared to 2019.^5^ A more detailed survey of mental health challenges undertaken in June 2020 by the CDC found that 40.9% of respondents had at least one adverse mental or behavioral health condition, including increased substance use and serious thoughts of suicide.^6^ These early reports collected data during the “lockdown period” of spring/summer 2020 at which time there were nearly universal stay-at-home orders, little was known about the novel coronavirus, and dramatic changes cut across social and occupational spheres.

In addition to surveillance efforts, research reports suggest that individuals with pre-existing mental illnesses may be particularly vulnerable to the mental health consequences of pandemic stressors.^7-9^ For example, a Dutch case-control cohort study found that the severity and chronicity of mental illness had a positive dose-response relationship with greater mental health burden and perceived mental health impact of COVID-19.^7^ A cross-sectional online study conducted in spring 2020 by Canadian and U.S. researchers found that participants with self-reported current anxiety disorders had higher levels of COVID-19-related stress than those with current mood disorders or those who did not report a current disorder.^8^ A longitudinal study conducted in Germany over a 12 week period in spring 2020 found similar results among those with pre-existing self-reported mental health diagnoses, namely higher mental health symptom levels in those with mood, anxiety, or other disorders.^9^

COVID-19 has persisted longer than expected, and research on mental health during the height of the pandemic is needed to better characterize the impact of associated psychosocial stressors, particularly among individuals with pre-existing mental health vulnerabilities. We describe a study, “The Mental Health Impact of COVID-19 Pandemic on NIMH Patients and Volunteers” (NCT04339790) launched in spring 2020 by researchers at the National Institute of Mental Health Intramural Research Program (NIMH IRP). For the study, we developed a new COVID-19-specific survey which captures two types of information, the psychological and behavioral responses to the pandemic, and the circumstances characterizing the pandemic period.

In this report, we utilized a subsample of well-characterized Patients and Controls to develop and validate a prediction model to estimate the mental health status for all study participants upon enrollment. We then evaluated whether the predicted probability of a participant being a “Patient” was associated with study measures of distress, loneliness, depression, several mental health factors, and COVID-19 related psychological and behavioral responses during the lockdown period (spring/summer 2020) in the U.S.

## METHODS

The study was carried out in accordance with the latest version of the Declaration of Helsinki and was approved by the Institutional Review Board of the National Institutes of Health. It represents a collaborative effort across several NIMH IRP labs and was rapidly developed in April 2020 in response to the emergent COVID-19 pandemic and anticipated mental health consequences. The study was conducted entirely online using a HIPAA compliant secure study website for electronic consent and completion of self-report questionnaires (nimhcovidstudy.ctss.nih.gov). Participants were asked to complete several measures every two weeks for six months.

The study sought to leverage existing clinical data from NIMH research participants, and thus recruitment began by inviting previously enrolled participants from several active NIMH IRP protocols. Shortly thereafter, a convenience sample from the general public was recruited via list-serv postings, social media ads, flyers, word of mouth, and the study listing on clinicaltrials.gov.

### Recruitment and NIMH subsample

The response to the study was rapid with nearly 6,000 requests for participation over the seven months of study enrollment (April-October 2020). Participants were considered enrolled if they completed the COVID-19 survey at the first study timepoint. Total study enrollment (n = 3,655) included participants from all 50 U.S. states and 40 international participants who entered the study from April 4 - November 1, 2020. The current paper focuses on data collected from 1,992 participants who enrolled during the first six weeks of the study (April 4–May 16, 2020). We focus on this “lockdown cohort” to control for ongoing changes in pandemic-related social and environmental stressors.

Within the lockdown cohort, more than 300 participants were previous NIMH IRP study participants across six labs. The previous participation was primarily in studies of mood and anxiety disorders; substance use disorders are exclusionary for most NIMH IRP studies. During this prior participation, 54.8% (174/317) of NIMH participants underwent a gold-standard psychiatric diagnostic interview (Structured Clinical Interview for DSM - SCID)^10^, and could thereby be classified as Patients (n=61) or Controls (n=113) based on lifetime history of a psychiatric disorder. On average, Patients were older and had a lower household income than Controls, so these demographic features were used as covariates in subsequent analyses. We will refer to this group of 174 participants as Patient/Controls.

### Measures

Study measures upon enrollment included: demographics, clinical history, functional status (WHODAS 2.0)^11^, psychiatric and family history (modified FIGS)^12^, alcohol (AUDIT)^13^and drug use (DSM-5 Level 2 Substance Use – Adult)^14^.

### Choice of primary measures

Primary self-report clinical outcome measures: distress (Kessler-5)^15^, loneliness (Three-Item Loneliness Scale)^16^, mental health symptoms (DSM-5 Self-Rated Level 1 Cross-Cutting Symptom Measure-Adult, (DSM-XC))^17^, and study-specific COVID-19-related responses and circumstances survey. These measures were completed at the enrollment time point and then every two weeks for six months. The current analysis uses data from the first (enrollment) time point only.

#### Distress

We used a five-item version of the Kessler High Distress Measure.^15^ The Kessler distress surveys have been used in population studies to screen for non-specific psychological distress and this version was used in Australia.^18^

#### Mental health symptoms

We chose the DSM-5 Level 2 Substance Use – Adult (DSM-XC) (https://www.psychiatry.org/psychiatrists/practice/dsm/educational-resources/assessment-measures)^17^ as one of the repeated measures due to its inclusion of transdiagnostic mental health symptoms. This measure is one of the core common data elements that NIMH researchers are expected to collect.^19^ Using data from all enrolled study participants, we performed a series of exploratory and confirmatory factor analyses and identified a six-factor solution which was robust across age, sex, and calendar time (see https://doi.org/10.1101/2021.04.28.21256253 ^20^). Each factor captured symptoms related to a general construct of psychopathology, which we provisionally termed: mood, worry, activation, somatic, thoughts, and substance use. Because two items from the Patient Health Questionnaire (PHQ-2) are embedded in the DSM XC, we could also calculate a depression score.^21, 22^

#### COVID-19 survey

We developed a self-report survey, the Psychosocial Impact of COVID-19 Survey, to capture a wide variety of COVID-19 related responses and circumstances (see <u>NIH Disaster Research Response)</u>. The survey was created in parallel with the CoRonavIruS Health Impact Survey (CRISIS) (crisissurvey.org) initiative led by other investigators within the NIMH IRP.^23, 24^ We implemented wording and content changes appropriate for adult respondents, including additional items (e.g., reported COVID-related physical symptoms, worries and expectations about the pandemic, and daily activities). Further, we embedded the three-item Loneliness Scale^16^ of perceived social isolation in this survey. Exploratory factor analyses did not support a coherent latent structure for the instrument (see ^25^), so with the exception of the Loneliness Scale, results are reported at the item level. The current analysis includes psychological and behavioral responses but not items about pandemic circumstances, e.g., financial problems, household composition.

### Data Analysis

The primary aim of our analysis was to evaluate the cross-sectional relationship between the mental health status classifier and a set of study outcome measures. In this section, we introduce the prediction model used to generate the classifier, its internal validation, and the subsequent analyses to relate the classifier to measures that reflect clinical symptoms and COVID-19 related psychological and behavioral responses.

#### Mental health status classifier

Using diagnoses obtained via the standardized psychiatric interview as the “ground truth” regarding mental health status in the Patient/Control group (n = 174), we developed a prediction for the binary variable of lifetime history of psychiatric disorder. The classifier takes as input the questions completed during the enrollment timepoint for the following surveys: demographics, self-reported psychiatric history for participant and family, functional status, alcohol use, and drug use.

We used a random forest classifier from the Python-based scikit-learn toolbox (maximum depth of 3, balanced class weighting)^26^. The classifier was evaluated first within the Patient/Control group (10-fold cross-validation, area under the curve of 0.92). We re-trained the classifier for all 174 Patient/Control participants, and then applied it to the full group of study participants in the lockdown cohort to generate a score based on their questionnaire data from enrollment. The classifier assigns a 0-1 probability of being a Patient, which we termed the “Patient Probability Score” (PPS). The ten most important survey items that contributed to the PPS are shown in Table 1.

**Table 1:** Contribution of Survey Items to Prediction Model.

| SURVEY ITEMS | IMPORTANCE |
| --- | --- |
| Have a diagnosis of depression?* | 0.12 |
| Feel very low for a couple of weeks or more?* | 0.08 |
| Have problems with nerves or emotions?* | 0.08 |
| How many days were these difficulties present?^ | 0.07 |
| Take medicine or see a doctor for [problems with nerves or emotions]? Take lithium?* | 0.07 |
| Concentrating on doing something for ten minutes?^ | 0.06 |
| How much of a problem did you have joining in community activities?^ | 0.06 |
| How many days did you cut back or reduce your usual activities or work?^ | 0.06 |
| Hospitalized for psychiatric problems?* | 0.03 |
| Attempt suicide?* | 0.03 |

\*Item from FIGS Survey (SELF)
^Item from WHODAS Survey

We next validated PPS against a composite indicator of mental health status derived from mental health treatment history on enrollment (1 for endorsement of mental health hospitalization, treatment for alcohol and/or drug abuse or treatment with medication for a mental health condition; 0 otherwise) (Table 2). This yielded an area under the curve of 0.87.

**Table 2.**
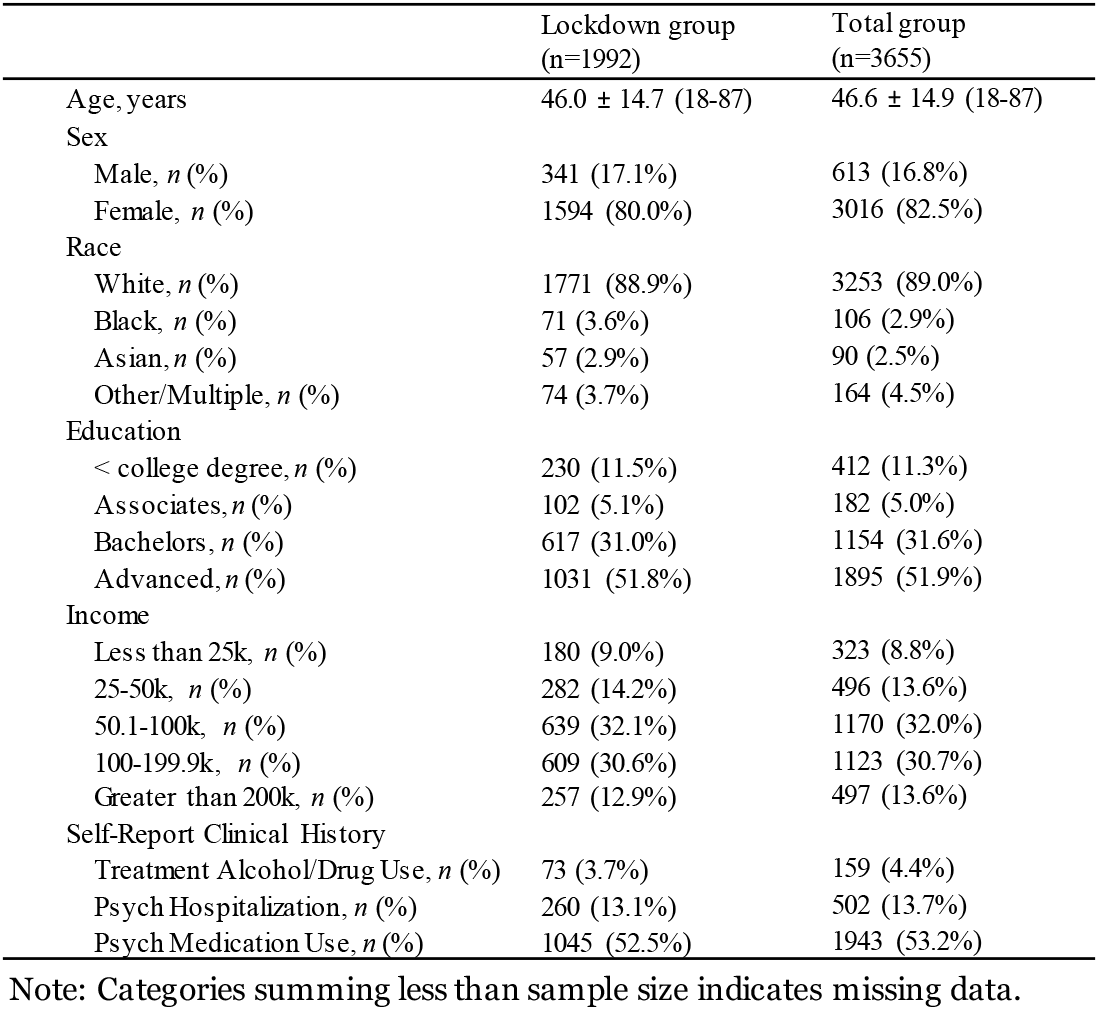
Demographics.

#### Relationship between Patient Probability Score and study outcomes

Regression models were used to analyze the cross-sectional relationship of predicted mental health status, operationalized by the PPS, and clinical measures (Kessler-5 total scores, 3-item Loneliness scores, PHQ-2 scores, DSM-XC factor scores), and COVID-19 survey psychological and behavioral responses at time of enrollment. Ordinary least squares regression was planned, but examination of model residuals indicated that the assumption of normality was not reasonable for the 3-Item Loneliness score, the PHQ-2 score, and the COVID-19 Survey items. Logistic regression was therefore used for these outcome measures. To accommodate the proportional odds assumption of ordinal logistic regression, the extreme response categories on some items were collapsed, and one item (change in exercise level) was binarized. Age and income were entered as covariates in all models. To improve interpretability, the PPS was multiplied by 10 such that a one-unit change reflected a 10% change on the 0 – 1 scale.

## RESULTS

### Demographics

The lockdown cohort (n=1,992) had a mean age of 46.0 (±14.7) years, was largely female (82.4%) and white (88.9 %) and did not differ in demographic profile from the full study sample (Table 2).

### PPS Associations with Study Measures of Clinical Symptoms and COVID-19 Related Responses

We leveraged diagnostic data from a subset of NIMH Patient/Controls to develop a prediction model that assigns a mental health status classifier, the Patient Probability Score (PPS), for each study participant. We then conducted analyses to relate the PPS to several concurrent clinical measures and COVID-19 related psychological and behavioral responses provided by participants during the lockdown period.

The PPS was positively correlated with all clinical measures: Kessler-5, Three-Item Loneliness, and PHQ-2 (Table 3), as well as each of the six DSM-XC mental health symptom factors (Figure 1). These scales and factors represent constructs of negative mental health outcomes such as distress, loneliness, and depression, that could reflect the impact of pandemic stressors. PPS was also more highly associated with the proposed DSM-XC factors of mood, worry, somatic and activation compared to thoughts and substance use factors.

**Table 3.** Regression Results for Association of PPS and Clinical Measures.

| **Linear Regression** | *n* | *B (SE)* | 95% CI | *t* |
| --- | --- | --- | --- | --- |
| Distress (K5) | 1992 | 1.049 (0.034) | [0.983, 1.115] | 31.15 |
| **Ordinal Logistic Regression** | *n* | *B (SE)* | *OR [95% CI]* | *t* |
| Loneliness (3-Item Scale) | 1979 | 0.258 (0.018) | 1.29 [1.25, 1.34] | 14.19 |
| Depression (PHQ-2) | 1984 | 0.550 (0.023) | 1.73 [1.66, 1.81] | 24.34 |

*Note:* SE = standard error; CI = confidence interval; OR= Odds Ratio; t = t statistic.

**Figure 1.**
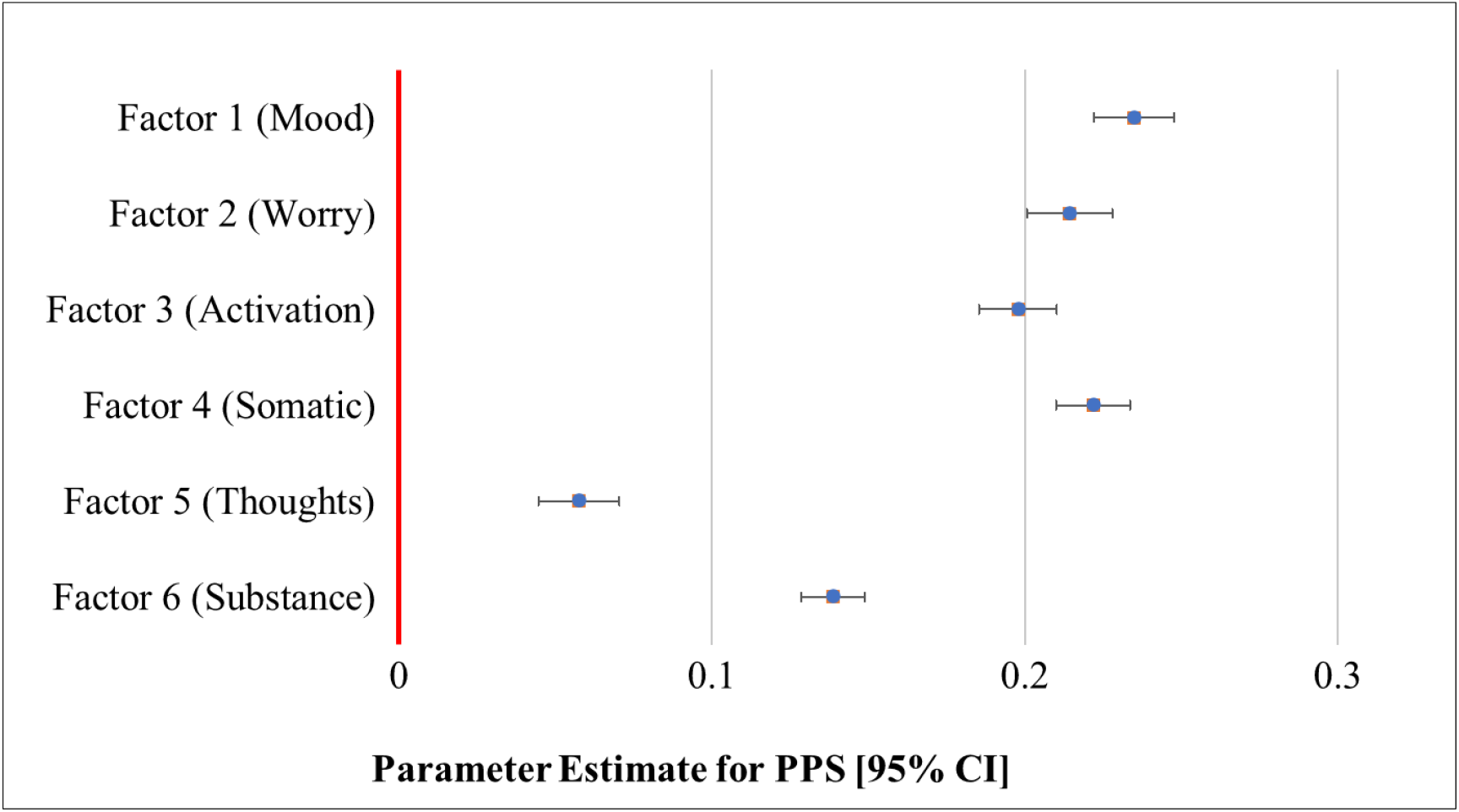
Linear Regression Parameter Estimates for PPS Associations with DSM-XC Factor Scores. *Note: D*ue to missing DSM-XC data, factor scores only computed on n = 1939.

Similarly, a higher PPS was associated with several different negative psychological responses on the COVID-19 survey such as worry, and lack of hope related to the pandemic (see Table 4). The largest effect of PPS was on worry that mental health could be affected by the pandemic, with the odds ratio demonstrating that for every 10% increase in PPS, the odds of worrying was multiplied by 1.46 [95% CI: 1.40, 1.51], followed by increased distress (OR 1.44 [95% CI: 1.39, 1.49]), and decreased ability to enjoy things (OR 1.35) or solve problems (OR 1.30). Worry about mental health exceeded worry about physical health (OR 1.13 [95% CI: 1.10, 1.17]) in our study population. PPS was also associated with several behavioral responses on the COVID-19 survey, including less time engaging in exercise, sleep, and mindfulness than usual. While PPS was not associated with adherence to social distancing guidelines, it was correlated with stress related to social distancing and worries about infection of self and others.

**Table 4.** Regression Results for PPS Association with COVID-19 Survey Responses.

| Items^ | *n* | *B (SE)* | *t* | *OR [95% CI]* |
| --- | --- | --- | --- | --- |
| **Psychological** |  |  |  |  |
| Able to concentrate/focus well | 1987 | -0.253 (0.019) | -13.55 | 0.78 [0.75, 0.81] |
| Hopeful coronavirus problem will end soon | 1979 | -0.132 (0.018) | -7.50 | 0.88 [0.85, 0.91] |
| Hope for a cure in the next year | 1981 | -0.077 (0.017) | -4.43 | 0.93 [0.89, 0.96] |
| Worry physical health could be affected by pandemic | 1974 | 0.127 (0.018) | 7.20 | 1.13 [1.10, 1.17] |
| Stressful maintaining social distancing | 1983 | 0.159 (0.018) | 8.97 | 1.17 [1.13, 1.21] |
| Worry self will be infected | 1982 | 0.154 (0.018) | 8.66 | 1.17 [1.13, 1.21] |
| Excessive need to seek information about COVID-19 | 1983 | 0.161 (0.018) | 9.10 | 1.17 [1.13, 1.22] |
| Worry others will be infected | 1985 | 0.172 (0.018) | 9.72 | 1.19 [1.15, 1.23] |
| Worry about coronavirus | 1984 | 0.200 (0.018) | 11.33 | 1.22 [1.18, 1.26] |
| Decreased ability to solve problems | 1975 | 0.264 (0.018) | 14.37 | 1.30 [1.26, 1.35] |
| Decreased ability to enjoy things | 1985 | 0.297 (0.018) | 16.20 | 1.35 [1.30, 1.40] |
| Increased distress during past week | 1983 | 0.364 (0.019) | 19.19 | 1.44 [1.39, 1.49] |
| Worry mental health could be affected by pandemic | 1975 | 0.375 (0.019) | 19.64 | 1.46 [1.40, 1.51] |
| **Behavioral** |  |  |  |  |
| Following social distancing guidelines | 1984 | 0.014 (0.019) | 0.72 | 1.01 [0.98, 1.05] |
| Change in time spent with others | 1985 | 0.016 (0.018) | 0.89 | 1.02 [0.98, 1.05] |
| Less time hobbies than usual | 1955 | 0.023 (0.018) | 1.29 | 1.02 [0.99, 1.05] |
| Avoiding information about COVID-19 | 1986 | 0.027 (0.017) | 1.52 | 1.03 [0.99, 1.06] |
| Change in physical activity | 1979 | 0.079 (0.017) | 4.52 | 1.08 [1.05, 1.12] |
| Increased time spent seeking COVID-19 information | 1975 | 0.091 (0.019) | 4.83 | 1.10 [1.06, 1.14] |
| Less time exercising than usual% | 1965 | 0.097 (0.020) | 4.77 | 1.10 [1.06, 1.15] |
| Less time mindfulness than usual | 1835 | 0.116 (0.019) | 6.24 | 1.12 [1.08, 1.16] |
| Less sleep than usual | 1971 | 0.156 (0.018) | 8.59 | 1.17 [1.13, 1.21] |

Note: Ordinal Logistic Regression Model used for all items except "Less time exercising". ; OR = odds ratio; CI = confidence interval; SE = standard error; t = t statistic; ^Item texted abbreviated in table. For full items, please see appendix. ; %Binary Logistic Regression model used

## DISCUSSION

There are broad and significant social and environmental stressors imposed by the COVID-19 pandemic on the public. Several studies have indicated that there may be differential impacts of pandemic stressors on those with pre-existing mental health conditions. Our convenience sample was recruited during a period of universal upheaval in the psychosocial and public health landscape (lockdown period). We found that mental health status, operationalized by PPS, was associated with concurrent clinical outcome ratings of distress, loneliness, depression, and mental health factor scores, as well as several COVID-specific psychological and behavioral responses. On average, associations between PPS and negative psychological responses were stronger than behavioral responses. PPS and specific COVID-19 survey item associations highlight pandemic-related psychological and behavioral concerns endorsed by those with mental health vulnerabilities during the period of lockdown.

Our study participants were not simply the “worried well.” A conservative estimate based on self-reported clinical history, finds that more than half of participants indicated a history of mental health treatment. However, we sought to better characterize mental health status based on participant responses to study measures at the time of enrollment, rather than rely on categorical grouping based on self-reported mental health history. Therefore, we applied machine learning methods to leverage existing clinical data on a subset of NIMH study participants with reliably diagnosed mental disorders. This led to the development of a continuous score of mental health status, which helps address a limitation of large-scale online studies for which full diagnostic assessments are not feasible.

## CONCLUSIONS

Our findings confirm that a focus on mental health during the COVID-19 pandemic is warranted, consistent with reports of an overall elevation in depressive and anxiety symptoms found early in the pandemic^6^. We show that these symptoms are accompanied by higher levels of pandemic-related negative psychological and behavioral responses among those with predicted mental health vulnerabilities. These associations should be followed over time as the COVID-19 pandemic and associated psychosocial factors, e.g., social unrest, are likely to change and evolve. We will include PPS when analyzing longitudinal trajectories of repeated study measures which will enable a more nuanced analysis of outcomes across the spectrum of participant mental health status. The use of a dimensional measure of mental health status when conducting the longitudinal analysis can better reveal the different trajectories among subgroups. In turn, these temporal patterns may identify risk factors associated with evidence of worsening clinical status, as well as resilience factors that may mitigate negative outcomes. Longitudinal analysis of study data will contribute needed information about how participants fared both at earlier stages of the pandemic and later, when circumstances differed.

## Data Availability

All data produced in the present study are available upon reasonable request to the authors.

## Limitations

The study recruited a convenience sample which does not reflect the demographic characteristics of those most affected by COVID-19 – lower income, racial and ethnic minorities. Online self-report responses cannot be verified and may be subject to social desirability bias. The patient sample from which the prediction model was derived did not include the full range of mental disorders, e.g., psychotic and substance use disorders.

## Data sharing statement

Deidentified data from the study will be made available and shared with others after publication through NIMH Data Archives (NDA).

